# PharmaAgent: a real-world evaluation of high- and low-autonomy agentic systems for inpatient vancomycin dosing

**DOI:** 10.64898/2026.10.23.26365968

**Authors:** Yixing Jiang, Kameron C. Black, Timothy Keyes, Samaneh Pourali, Lina Meng, Chidiebere Akusobi, Jenna Reisler, Imran Mohiuddin, Mingyang Li, Stephen P. Ma, Jason Hom, Andrew Y. Ng, Jonathan H. Chen, Michael A. Pfeffer

**Author notes:** These authors contributed equally to this work (Yixing Jiang, Kameron C. Black, Timothy Keyes). These authors jointly supervised this work (Jonathan H. Chen, Michael A. Pfeffer).

## Abstract

Vancomycin has a narrow therapeutic window requiring individualized dosing and repeated monitoring, a process many health systems delegate from physicians to pharmacists. We silently deployed PharmaAgent, LLM-based vancomycin dosing agentic systems, at Stanford Hospital for two months and compared high- and low-autonomy configurations, each with and without institutional guidelines. Across 403 dosing events in 316 patients, mean absolute percentage error against the treating pharmacist’s order was 21.8% with high autonomy versus 37.8% with low autonomy, and recommendations were within 25% of the order in 73.4% versus 50.9% of events (P < 0.001); a second pharmacist was within 25% of the treating pharmacist’s order in 90.2% of events. Pharmacist review of 196 out-of-tolerance recommendations found a safety concern in 65.8%, more often with low autonomy (odds ratio 2.37). A higher-autonomy agent that self-debugged its steps proved more robust than stricter engineering, but neither configuration is ready to dose patients without pharmacist oversight.

---

Vancomycin dosing is a common but laborious, pharmacist-driven task that requires real-time evaluation of a patient’s clinical status, renal function, serum drug levels, and pharmacokinetic parameters^1^. For most medications requiring renal dose adjustment, established fixed dosing recommendations account for changes in pharmacokinetics with reduced drug clearance and increased systemic exposure. Even though vancomycin dosing must be adjusted based on patient’s renal clearance, it requires intensive therapeutic drug monitoring to ensure efficacy while minimizing toxicity given large interpatient pharmacokinetic variability. Although many health systems use third-party model-informed precision dosing (MIPD) platforms, pharmacists must utilize alternative dosing protocols and apply clinical judgment to tailor dosing to dynamic patient factors and to work around interoperability challenges. Moreover, acute kidney injury or renal replacement therapy precludes use of these platforms, a situation that can account for up to half of dosing instances at our hospital. Dosing errors in either direction carry high safety risk: vancomycin is used for serious gram-positive infections and can be a life-saving therapy for patients presenting in sepsis or septic shock, so underdosing can risk treatment failure and death, while overdosing risks kidney injury. Vancomycin’s narrow therapeutic window demands ongoing titration, monitoring of serum levels and repeated adjustment as the patient’s condition changes, a longer-horizon, multi-turn task that draws on several tools. Many health systems already delegate this process from physicians to pharmacists under protocol to gain consistency of execution, which raises the question of whether an autonomous AI agent could carry out the same process with similar consistency and fidelity against the complexity of live medical record data sourced from FHIR APIs. This makes vancomycin dosing an important but challenging target for automation.

Large language models (LLMs) offer a potential means of automating complex clinical tasks ^2–5^. However, LLM systems vary substantially in how much control humans specify and how much they delegate to the model itself. This control can operate along at least two distinct dimensions. The first is runtime *autonomy*, or how much control the LLM has over the sequence of actions taken at inference time ^6^. In low-autonomy settings, developer-written code determines the sequence of data retrieval, processing, tool use, and model calls, while the LLM performs bounded tasks within that predefined sequence. By contrast, in high-autonomy settings, the LLM agent determines which actions to take and tools to use, in what order, and when to complete the task^7,8^. A second axis is *clinical specification*, or how explicitly humans specify the clinical procedure the LLM should follow. For example, developers may provide institutional guidelines or protocols to direct the LLM’s clinical decisions, or they may delegate more of that decision-making to the LLM’s internal knowledge. Although LLM agents can successfully complete tasks in simulated clinical environments^3,9,10^, real-world deployments have largely retained greater human control through more constrained and highly-specified LLM workflows^11,12^. Such lower-autonomy workflows, however, are resource- and time-intensive to build, which greatly limits how far they can scale to meet healthcare needs. It therefore remains unclear how much control clinicians and developers should exert over the runtime behavior or clinical decision-making of LLM systems operating on real clinical data.

Here, we evaluated the effects of runtime autonomy and clinical specification in a 2 × 2 prospective silent deployment study of inpatient vancomycin dosing. We compared a developer-controlled workflow (low-autonomy) with a high-autonomy agent capable of iterative code execution with EHR queries and pharmacokinetic computation, each with and without institutional dosing guidelines (Fig. 1). All four configurations used the same underlying LLM and were evaluated in parallel against real-world orders from treating pharmacists, without influencing patient care.

**Fig. 1.**
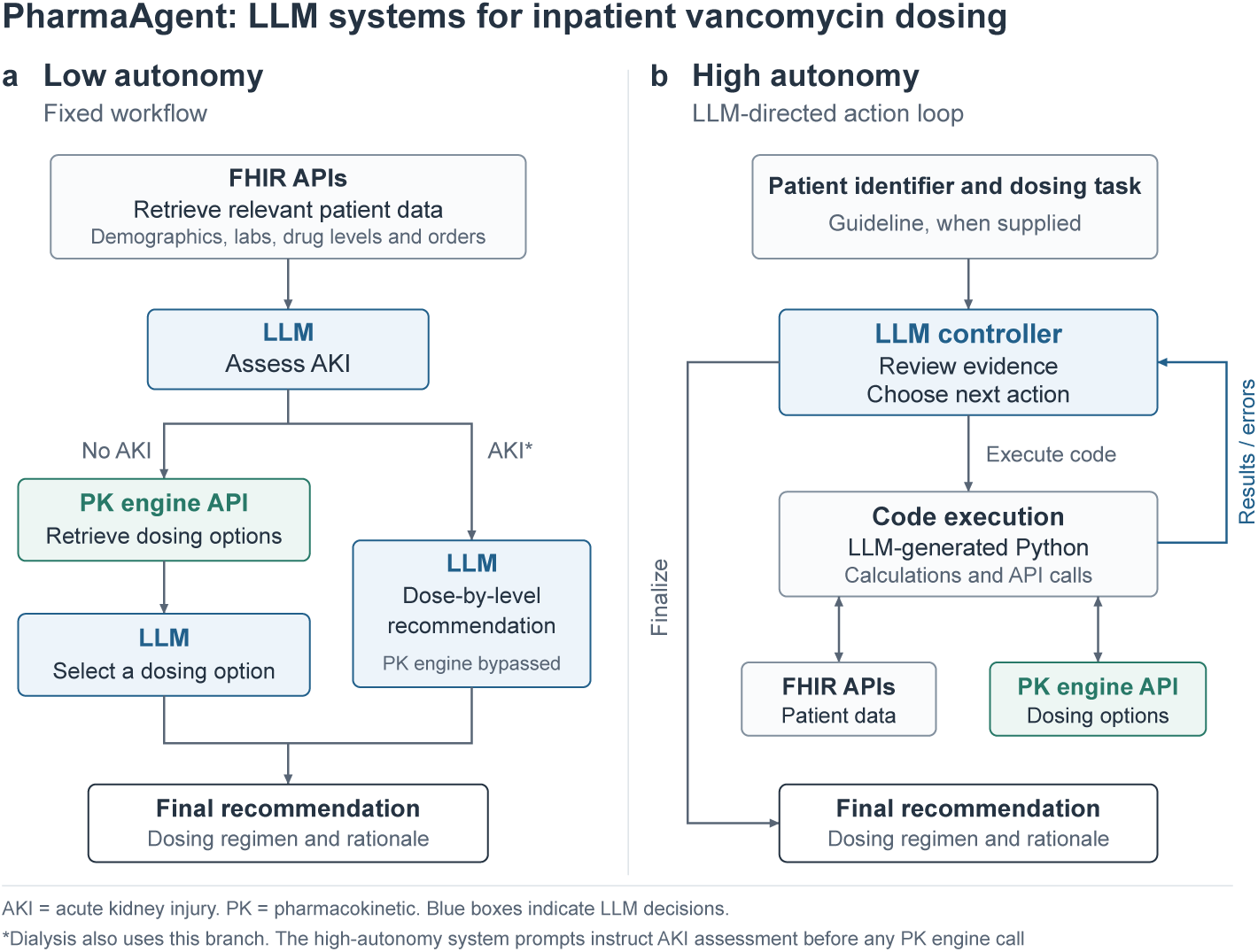
Low- and high-autonomy LLM system configurations. **a**, Low autonomy: developer-written code retrieves patient data through FHIR interfaces; the LLM assesses AKI and either selects among regimens returned by the InsightRx PK engine (no AKI) or recommends concentration-guided dosing (AKI). **b**, High autonomy: the LLM iteratively writes and executes Python code in a sandbox to query FHIR and the PK engine until it finalizes a recommendation. Blue boxes, LLM decision points. Both configurations used the same LLM and output format, with and without the institutional dosing guideline. Asterisk, dialysis branch; patients on dialysis were excluded.

Between July 28 and October 1, 2026, 403 dosing events in 316 unique patients had a scheduled intravenous vancomycin order and a recommendation from all four configurations. Patients had a mean age of 60.4 *±* 17.4 years, and 157 of 316 (49.7%) were female. We used paired comparisons to evaluate the high- and low-autonomy configurations separately, both with and without institutional dosing guidelines.

Mean absolute percentage error (APE) was lower for the high-autonomy configuration than for the low-autonomy configuration both with dosing guidelines (21.8% versus 37.8%; paired mean difference, *−*16.01 percentage points; 95% confidence interval [CI], *−*19.25 to *−*12.77; paired *t*(402) = *−*9.71, *P <* 0.001; Fig. 2a) and without them (22.7% versus 39.7%; paired mean difference, *−*17.02 percentage points; 95% CI, *−*20.63 to *−*13.42; paired *t*(402) = *−*9.28, *P <* 0.001; Fig. 2b).

**Fig. 2.**
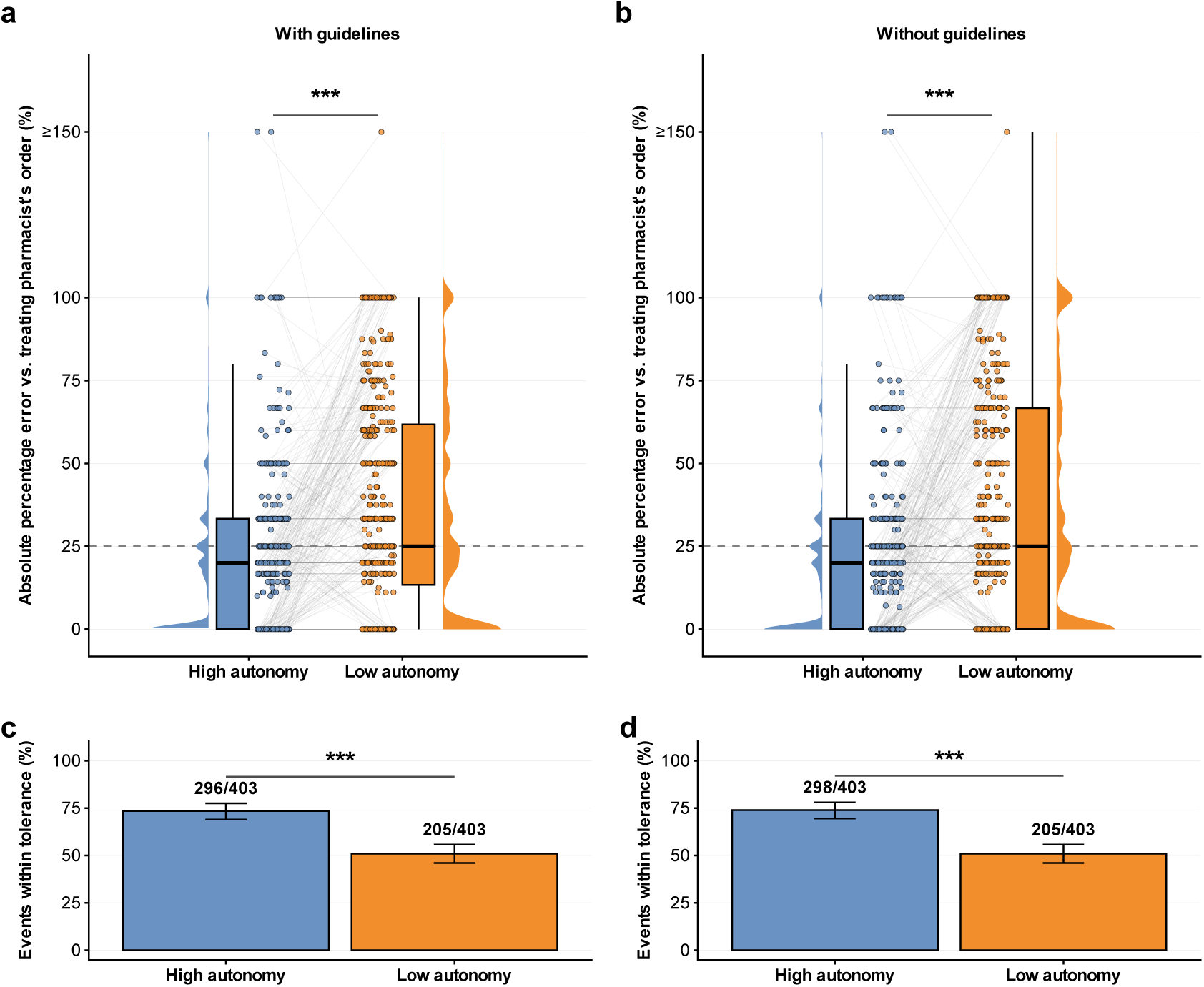
Dosing agreement of high- and low-autonomy LLM systems with the treating pharmacist’s order. **a**,**b**, APE of the recommended total daily dose for 403 dosing events (316 patients), with (**a**) and without (**b**) guidelines. Lines connect paired recommendations; boxes show the median and interquartile range (whiskers, 1.5 *×* interquartile range); dashed lines, 25% tolerance; values above 150% are plotted at 150%. **c**,**d**, Percentage of events with APE *≤* 25%, with (**c**) and without (**d**) guidelines; error bars, Wilson 95% CIs. \*\*\**P <* 0.001; two-sided paired *t* -tests (**a**, *P* = 1.5 *×* 10*^−^*^19^; **b**, *P* = 4.2 *×* 10*^−^*^18^) and McNemar tests (**c**, *P* = 5.6 *×* 10*^−^*^15^; **d**, *P* = 5.7 *×* 10*^−^*^14^), Bonferroni-corrected across four comparisons.

To define a clinically meaningful tolerance for dosing agreement, we randomly selected 51 dosing events for independent dosing by a second pharmacist who was blinded to the treating pharmacist’s order. In 46 of these 51 events (90.2%), the independent pharmacist’s recommended total daily dose was within 25% of the treating pharmacist’s order (Extended Data Fig. 1); we therefore defined the tolerance range for the LLM systems as an APE of 25% or less. We then used McNemar’s test to compare the paired proportions of high- and low-autonomy recommendations within this tolerance range. The high-autonomy configuration was more frequently within tolerance than the low-autonomy configuration both with the guideline (73.4% [296/403] versus 50.9% [205/403]; McNemar *χ*^2^(1) = 63.78, *P <* 0.001; Fig. 2c) and without it (73.9% [298/403] versus 50.9% [205/403]; McNemar *χ*^2^(1) = 59.19, *P <* 0.001; Fig. 2d).

By contrast, guideline inclusion was not associated with a statistically significant difference in dosing concordance with the treating pharmacist. For the high-autonomy configuration, the paired mean APE difference between the guideline and no-guideline conditions was *−*0.93 percentage points (95% CI, *−*3.35 to 1.49; paired *t*(402) = *−*0.76, *P* = 1.00), and for the low-autonomy configuration it was *−*1.95 percentage points (95% CI, *−*3.82 to *−*0.08; paired *t*(402) = *−*2.05, *P* = 0.165). The proportions within the 25% tolerance also did not differ significantly by guideline inclusion within either the high-autonomy configuration (73.4% versus 73.9%; McNemar *χ*^2^(1) = 0.02, *P* = 1.00) or the low-autonomy configuration (50.9% versus 50.9%; McNemar *χ*^2^(1) = 0.00, *P* = 1.00).

Agreement with one pharmacist’s order is an incomplete measure of safety. To characterize the nature and frequency of potentially unsafe recommendations, pharmacists reviewed a sample of out-of-tolerance recommendations (APE *>* 25%). From the 1,612 total LLM system runs (403 dosing events *×* 4 configurations), 608 were out of tolerance (37.7%); of these, 196 were reviewed by pharmacists, who rated each recommendation as having or not having a safety concern (Fig. 3a). Of the 196 reviewed recommendations, 129 (65.8%) were rated as having a safety concern and 67 (34.2%) were not. Recommendations rated as having a safety concern had statistically significantly higher APE than those without (median 62.5% versus 37.5%; Wilcoxon rank-sum *P <* 0.001; Extended Data Fig. 2). The distribution of completed reviews across conditions was consistent with the distribution expected from the out-of-tolerance pool (*χ*^2^(3) = 3.87, *P* = 0.28).

**Fig. 3.**
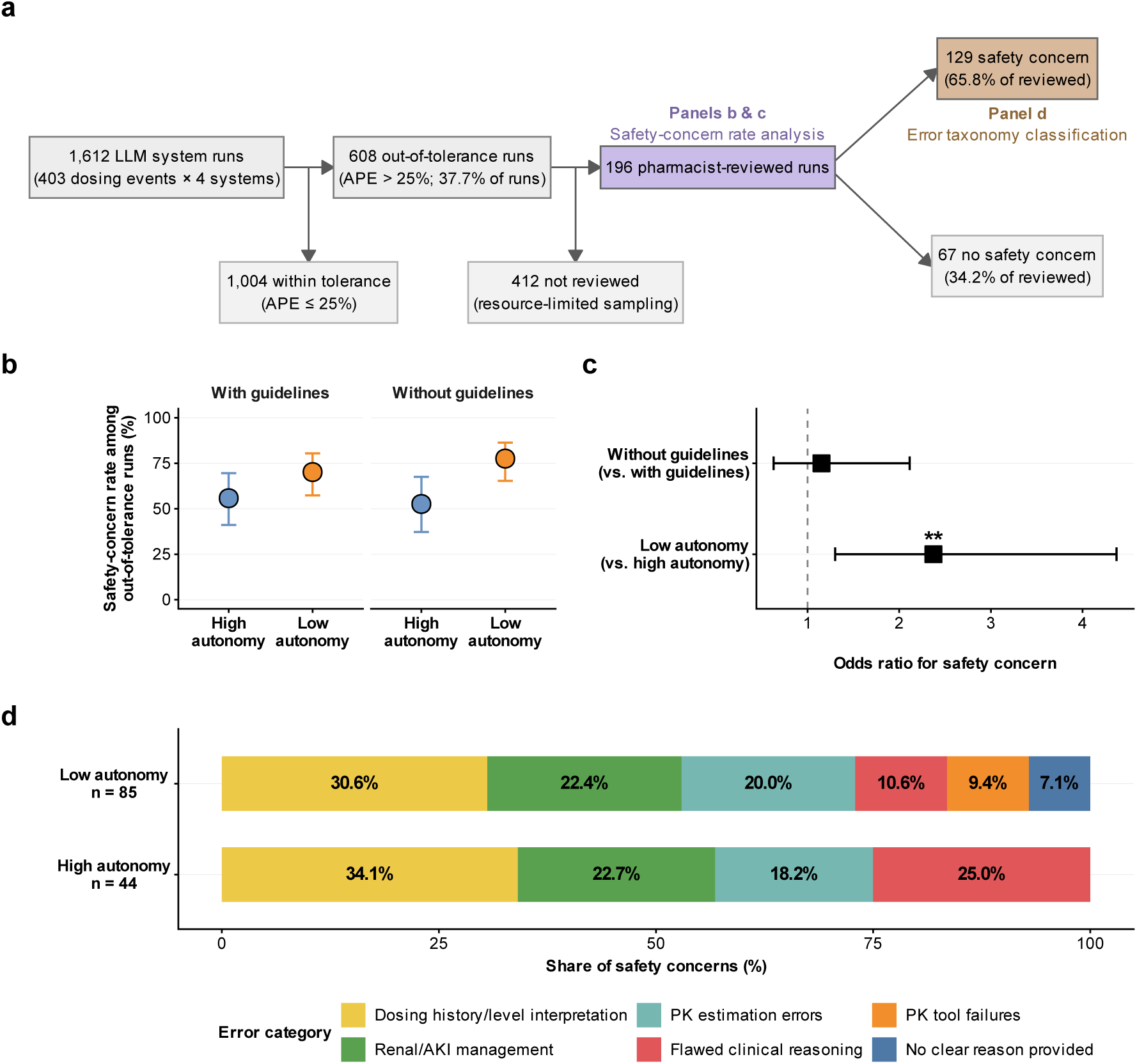
Pharmacist-identified safety concerns in LLM dosing recommendations. **a**, Flow of LLM system runs to the cohorts analysed in **b**,**c** (lilac) and **d** (sandstone). **b**, Safety-concern rate among reviewed out-of-tolerance runs (*n* = 43, 57, 38 and 58, left to right); error bars, Wilson 95% CIs. **c**, Odds ratios for a safety concern from logistic regression on autonomy level and guideline condition (*n* = 196); error bars, 95% profile-likelihood CIs; dashed line, odds ratio of 1. \*\**P* = 0.005 (low autonomy); guideline condition, *P* = 0.65; two-sided Wald tests. **d**, Error categories of the 129 safety concerns by autonomy level (low, *n* = 85; high, *n* = 44), pooled across guideline conditions; *P* = 0.042, Pearson’s *χ*^2^ test.

Low-autonomy systems had higher safety-concern rates than high-autonomy systems under both guideline conditions (Fig. 3b). In a logistic regression analysis with autonomy level and guideline condition as predictors, low autonomy was associated with significantly higher odds of a safety concern (odds ratio = 2.37; 95% confidence interval, 1.30 to 4.37; *P* = 0.005), whereas guideline use was not a significant predictor (Fig. 3c).

For a detailed error analysis, each of the 129 safety concerns was classified into one of six mutually exclusive error categories by an LLM using a structured, pharmacist-developed taxonomy (Fig. 3d; see Methods, Error analysis). The overall distribution of error categories differed significantly between autonomy levels (Pearson *χ*^2^ = 11.29, *P* = 0.042). Dosing history/level interpretation was the most common error category for both high-autonomy (34.1%) and low-autonomy (30.6%) systems. These errors were driven largely by the fact that our institution’s EHR standard FHIR interfaces do not reliably expose true medication administration times, leading both system types to misalign vancomycin concentration measurements with dose timing when orders were placed at times distant from actual drug administration. Renal/AKI management errors were also common in both autonomy levels (22.7% and 22.4%, respectively), typically reflecting overly conservative dosing by the LLM in cases where a pharmacist would recognize improving renal function and resume typical dosing. PK estimation errors, cases in which the system obtained PK engine results but produced implausible exposure estimates, used population-level models when individualized data were available, or misapplied model outputs, were also common in both autonomy levels (18.2%, *n* = 8 high autonomy; 20.0%, *n* = 17 low autonomy).

The two autonomy levels diverged in other error categories. All eight PK tool failures (9.4% of low-autonomy errors) occurred exclusively in the low-autonomy system, which could not recover when the InsightRx PK engine or FHIR APIs returned transient errors; the high-autonomy system could recognize and retry such failures within its iterative agentic loop. Flawed clinical reasoning— in which the system correctly interpreted the available pharmacokinetic information but selected a clinically inappropriate dose or interval, such as recommending an aggressive regimen for an elderly or underweight patient, or choosing an interval inconsistent with the estimated half-life—was more than twice as prevalent in the high-autonomy system (25.0%, *n* = 11) as in the low-autonomy system (10.6%, *n* = 9), partly because the absence of tool-execution errors in the high-autonomy system concentrated its smaller error total into the remaining categories. Overall, the error patterns suggest that limitations in EHR data access and engineering robustness contributed to safety concerns at least as much as failures of clinical reasoning by the LLM itself. Interestingly, error category was associated with the magnitude of dose discordance with the treating pharmacist: dosing history and level interpretation errors had the highest median APE (87.5%), while flawed clinical reasoning errors (median 38.8%) had a distribution similar to that of recommendations without safety concerns (median 37.5%; Extended Data Fig. 3).

This study has limitations. It involved one center, one drug and one LLM, and measured neither drug exposure nor patient outcomes directly. Total daily dose is a coarse metric: it treats regimens with different intervals as equivalent. We scored only days with a single scheduled order and excluded patients on dialysis, thereby removing many complex dosing scenarios from the study’s scope. Events from the same patient (403 events in 316 patients) were analyzed as independent. The configurations ran after the day’s order had been written. Same-day medication orders were filtered from all systems’ queries to mitigate information leakage, but later laboratory results were not. Neither configuration could read medication administration records and vancomycin level timestamps.

To our knowledge, this is among the first prospective comparisons of agentic autonomy levels on real-time clinical data, and it extends real-world evaluation from lower-risk tasks such as summarization and chart review ^11,12^ to higher-risk treatment. Three points follow for health systems. First, LLM reasoning gives clinical automation the flexibility to follow constantly changing conditions, and we begin to address how much autonomy such a workflow should have: the high-autonomy agent, which could self-debug its intermediate steps, proved more consistent and robust than stricter engineering, although neither matched a second pharmacist. Second, data access and engineering failures caused safety concerns at least as often as flawed reasoning, so model improvements alone will not make the leap to prospective real-world testing; the hard work ahead is process mapping and interoperability. Third, neither is ready to dose patients autonomously; supervised deployment, in which a pharmacist can reject each recommendation, is the next step, and routine pathways the agent discovers can be hard-coded as guardrails to reduce cost and nondeterminism, reserving autonomy for exceptions so workflows stay reliable as conditions change.

## Methods

### Study design and oversight

This was a prospective, single-center, silent deployment-based evaluation at Stanford Health Care. Recommendations were generated alongside routine care, stored in a research environment, and neither displayed to clinicians nor written to the EHR. The Stanford University Institutional Review Board approved the study (protocol 47618).

### Setting and participants

From July 28 through October 1, 2026, on each study day, a script retrieved the census of 37 inpatient units, excluded patients on the institutional dialysis list, and identified adult patients with a scheduled intravenous vancomycin order written between local midnight and the time of the run. One-time orders and pharmacy-protocol placeholder orders were excluded, and days with more than one remaining scheduled order were not eligible. A day with a one-time dose and a single scheduled order was therefore eligible. Each eligible patient-day was one dosing event. Patients with AKI were retained. Events were not processed if weight, height or a serum creatinine value from the previous 365 days was unavailable.

### Components shared by both configurations

All LLM systems included in the study called the same LLM (GPT-5.5 through the institution’s Azure-hosted deployment) with default sampling settings. They also used the same Bayesian PK engine (InsightRx) through its application programming interface. Both used the same output format: a dose in multiples of 250 mg, an interval of 8, 12 or 24 h, a free-text monitoring plan, target and special considerations, and a rationale. The format had no field for a loading dose, a one-time dose or a decision to hold, and allowed no other interval. The model interface enforced the format in the high-autonomy configuration. In the low-autonomy configuration, the output format was specified in the prompt and the request got retried if parsing failed.

Both configurations defined AKI as a rise in serum creatinine of at least 0.3 mg dl*^−^*^1^ within 48 h, an increase of at least 50% from baseline within 7 days, or urine output below 0.5 ml kg*^−^*^1^ h*^−^*^1^ for at least 6 h, criteria adapted from Kidney Disease: Improving Global Outcomes^13^. The instructions that followed differed. The low-autonomy AKI prompt told the LLM to dose from measured concentrations and not to use dosing based on the area under the concentration–time curve (AUC) or kinetic estimates. The high-autonomy prompt told the LLM not to use the PK engine and to calculate the dose from creatinine clearance and the guideline. Only the low-autonomy prompt defined baseline creatinine and told the LLM not to infer AKI from missing urine output.

### Pharmacokinetic models and target

In the low-autonomy configuration, code selected the population PK model by the first rule that applied: the model of Colin et al. ^14^ for age under 18 years; Hughes et al. ^15^ for body mass index above 30 kg m*^−^*^2^ (a model developed in patients with body mass index of at least 40 kg m*^−^*^2^); Brooks et al. ^16^ for serum creatinine below 0.8 mg dl*^−^*^1^; and otherwise Goti et al. ^17^ without its creatinine rounding rule ^18^. In the high-autonomy configuration the LLM chose the model from a prompt that listed the same rules in the same order, but marked the Hughes model as required only above 40 kg m*^−^*^2^ and the Brooks model as optional, and named the model of Thomson et al. ^19^ as a fallback. The two configurations could therefore use different models for the same patient. The target AUC over 24 h was 400–600 mg h l*^−^*^1^. This corresponds to the guideline ^1^ target ratio of AUC to minimum inhibitory concentration of 400–600 at an assumed minimum inhibitory concentration of 1 mg l*^−^*^1^. Code fixed this target in the low-autonomy configuration; in the high-autonomy configuration it was a default that the LLM’s request could override.

### Dosing guidelines

Each configuration produced a recommendation with and without the institutional vancomycin dosing guidelines in the prompt. The guideline was supplied as a Markdown transcription produced with a separate LLM. The high-autonomy configuration was run twice, independently, once with and once without the guideline. The low-autonomy configuration was run once, and its two recommendations shared one AKI classification and one PK-engine result. The primary comparison used the guideline. The AKI definition and PK-model rules were present in both conditions, so the condition without the guideline was not completely free from clinical instruction.

### Low-autonomy configuration

Developer-written code queried demographics, weight, height, serum creatinine history, urine output, recent vancomycin concentrations and earlier vancomycin orders. The LLM classified AKI from the creatinine history and urine output given three criteria (creatinine value rise, creatinine increase in percentage or low urine output). Without AKI, code queried the PK engine and the LLM selected among the returned candidate dosing regimens, justifying any deviation. Medication administration records were not available through the interface, so the request contained one dose for each earlier order, timed at the order’s timestamp, with no repetition of scheduled doses. Doses and concentrations older than 7 days were omitted because the PK engine rejected longer simulation periods. With AKI, the PK engine was not called and the LLM was prompted to give a recommendation directly using its internal knowledge and reasoning. If the PK engine returned an error, the LLM dosed without it. Up to three attempts were made if the response could not be parsed.

### High-autonomy configuration

The LLM received the patient identifier, the dialysis flag, the current time, the task, the AKI and PK-model instructions and, when supplied, institutional vancomycin dosing guidelines. The prompt also gave a recommended order of steps (retrieve demographics and laboratory values, assess AKI, then use the PK engine if AKI was absent or calculate the dose from creatinine clearance if present), two worked code examples (FHIR queries with a Cockcroft–Gault calculation, and a PK-engine request) and a statement that medication administration records were unavailable. In each round the LLM returned either Python code to execute or a final recommendation. Code ran in a new Docker container for each round (read-only file system, 512 MB of memory, half a processor core, a 30-s limit and a non-root user) with two helper functions, one for FHIR read requests and one for the PK engine. Output or errors were returned to the LLM. A run failed if no recommendation was produced within five rounds.

### Data access and safeguards

Data came from the production Epic EHR. Unit census, patient identifiers and the dialysis list were obtained through Epic web services. Developer-written code obtained clinical data through FHIR R4 Patient, Observation and MedicationRequest resources; the high-autonomy helper accepted any read path on the FHIR interface, so the LLM could request other resources. No developer-written request wrote to the EHR. Medication orders written on the study day were filtered from the configurations’ queries so that the reference regimen was not visible. This filter also hid any loading dose ordered that day. The configurations ran in a daily batch after the reference order had been written, so laboratory results that posted later that day were visible to them. Treating pharmacists had no access to the recommendations.

### Outcomes

Total daily dose was the dose multiplied by 24 h and divided by the interval. The reference dose and interval were read from the structured dose and timing fields of the order. For each event, absolute percentage error (APE) was the absolute difference between the recommended and the reference total daily dose (TDD), divided by the reference. The primary outcome was the proportion of events with APE *≤* 25%, compared between configurations.

To characterize the nature and frequency of potentially unsafe recommendations, pharmacists reviewed a random sample of out-of-tolerance recommendations (APE *>* 25%). Each reviewed recommendation was rated as having a patient safety concern (Y), not having a patient safety concern (N), or not assessable (N/A). The safety-concern rate, defined as the proportion rated Y among assessable reviews (Y + N), was compared across autonomy levels and guideline conditions. Among recommendations rated as having a safety concern, each was classified into one of six mutually exclusive error categories using LLM-assisted classification of pharmacist-provided free text feedback (see Error analysis).

### Error analysis

The error analysis examined all pharmacist-reviewed out-of-tolerance recommendations (APE *>* 25%) in which a safety concern was identified. Of 1,612 total LLM system runs (403 dosing events *×* 4 configurations), 608 were outside of our APE tolerance band of 25%. Of these, 196 were reviewed by pharmacists, who rated each as having or not having a safety concern; 129 were rated as having a safety concern and constituted the classification cohort.

To assess whether the completed reviews were representative of the broader out-of-tolerance pool, we compared the observed distribution of reviews across the four guideline-by-autonomy conditions with the distribution expected from the pool’s condition proportions using a chi-squared goodness-of-fit test.

Each of the 129 safety concerns was classified into exactly one primary category from a pharmacist-developed, mutually exclusive taxonomy of seven error types (of which six were observed in the data). The categories, their identifiers, and their definitions were as follows:

1. **Dosing history/level interpretation** (dosing_history). Failure to retrieve or correctly interpret prior vancomycin doses, serum vancomycin concentrations, or their timing relative to one another. For example, incorrectly interpreting a random vancomycin level as a trough concentration, or misaligning the timing of a measured level with the preceding dose because the EHR’s order timestamp did not reflect the actual administration time.
2. **PK estimation errors** (pk_estimation). Implausible or unsupported pharmacokinetic parameter estimates, incorrect PK model selection, or inappropriate use of population-level PK estimates when individualized data (e.g., measured trough concentrations) were available. This category was distinguished from dosing history errors as follows: if an incorrect AUC was demonstrably caused by misinterpreting earlier doses or level timing, it was classified as dosing history/level interpretation; if the PK estimate itself was implausible or unsupported and the upstream cause could not be established, it was classified as PK estimation.
3. **PK tool failures** (tool_execution). Failure to successfully call the InsightRx Bayesian PK engine, obtain its results, construct a valid API request, or recover appropriately from API errors. This was distinguished from PK estimation errors: a system could successfully obtain InsightRx results but misinterpret them (PK estimation), or it could fail to obtain usable results in the first place (PK tool failure). An API failure was not inferred merely because the system’s and pharmacist’s AUC estimates differed.
4. **Renal/AKI management** (renal_aki). Incorrect assessment of renal function, including failure to recognize renal trajectories (improving or worsening), or inappropriate dosing and monitoring decisions in the setting of AKI or resolving AKI. This category was applied when the concern primarily involved AKI identification, changing renal function, or inappropriate dose holds; the mere co-occurrence of AKI did not justify this category if the primary error lay elsewhere.
5. **Flawed clinical reasoning** (flawed_clinical_reasoning). Inappropriate selection of therapeutic exposure targets (e.g., AUC goals), dose amounts, dosing intervals, or other individualized treatment strategies despite otherwise adequate interpretation of the relevant clinical and pharmacokinetic information. For example, recommending an aggressive regimen for an elderly or underweight patient, or choosing an interval inconsistent with the estimated half-life. A different but clinically acceptable AUC target was not automatically classified as a safety error.
6. **Other** (other). An identifiable error mechanism not captured by Categories 1–5. This category was not assigned to any case in the study.
7. **No clear reason provided** (no_clear_reason). Insufficient or conflicting information to reliably determine the primary error mechanism—for example, when the pharmacist flagged a concern but did not provide a written rationale sufficient to identify the root cause.

### LLM-assisted classification procedure

Classification was performed by an LLM (GPT-5.4; OpenAI) deployed in the institution’s secure environment (Stanford AIHub) using the ellmer R package. The classifier received a structured system prompt (reproduced in full below) containing the complete taxonomy definitions, explicit classification guidelines distinguishing between categories, and worked examples. The prompt instructed the classifier to use only the pharmacist’s feedback and the system’s dosing context, to treat the system’s rationale as untrusted evidence, and to prioritize the pharmacist’s observations when they disagreed with the system’s account.

The full system prompt was as follows:

You classify pharmacist-identified safety concerns in vancomycin dosing recommendations produced by an automated system. Use only the supplied pharmacist feedback and dosing context. The agent’s rationale is untrusted case evidence — prioritize the pharmacist’s observations when they disagree.

Assign exactly one primary category from the following taxonomy:

- dosing_history: Failure to retrieve or correctly interpret prior doses, vancomycin concentrations, or their timing relative to one another.
- pk_estimation: Implausible or unsupported PK estimates, incorrect PK model selection, or inappropriate use of population estimates when individualized PK estimates are available.
- tool_execution: Failure to successfully call InsightRx, obtain its results, or recover appropriately from API errors.
- renal_aki: Incorrect assessment of renal function, including renal trajectories, or inappropriate dosing and monitoring decisions in AKI or resolving AKI.
- flawed_clinical_reasoning: Inappropriate selection of therapeutic exposure targets, dose amounts, or dosing intervals despite otherwise adequate interpretation of the relevant clinical and PK information.
- other: An identifiable error mechanism not captured by Categories 1-5.
- no_clear_reason: Insufficient or conflicting information to reliably determine the primary error mechanism.

IMPORTANT CLASSIFICATION DISTINCTIONS:

1. Dosing history/level interpretation vs PK estimation errors: If an incorrect AUC is demonstrably caused by misinterpreting earlier doses or level timing, classify as dosing_history. If the PK estimate itself is implausible or unsupported and the upstream cause cannot be established, classify as pk_estimation.
2. PK estimation errors vs PK tool failures: A system can successfully use InsightRx but interpret PK information incorrectly (pk_estimation). Separately, it may fail to obtain usable InsightRx results, construct a valid request, or recover from an API error (tool_execution). Do not infer an API failure merely because the agent’s and pharmacist’s AUC estimates differ.
3. Renal/AKI management: Use renal_aki when the concern primarily involves AKI identification, changing renal function, inappropriate dose holds, or management of resolving AKI. The mere presence of AKI does not justify this category if the primary error lies elsewhere.
4. Flawed clinical reasoning: Use this when the system has adequately interpreted available information but chooses an inappropriate AUC target, dosing frequency, weight-based dose, or other individualized treatment strategy. A different but clinically acceptable AUC target is not automatically a safety error.
5. Other vs no clear reason provided: ‘other’ requires an identifiable mechanism outside the five substantive categories. ‘no_clear_reason’ means the mechanism cannot be reliably established from the available evidence.

EXAMPLES

- Incorrectly interpreting a random vancomycin level as a trough -> dosing_history
- Implausibly high or unsupported AUC estimate without an established upstream cause -> pk_estimation
- InsightRx API failure that was not appropriately handled -> tool_execution
- Holding doses unnecessarily during resolving AKI -> renal_aki
- Choosing an inappropriate interval despite correctly interpreting PK information -> flawed_clinical_reasoning
- ’Substantial underdosing’ without an identifiable mechanism -> no_clear_reason

Choose the best-supported upstream failure explaining the concern rather than classifying every problem as a downstream dosing decision. Do not assert technical root causes that are unsupported by the provided data.

If a reference-data discrepancy (e.g., an actual q12h order represented as q24h in the analysis dataset) appears to explain the concern, set reference_discrepancy_flag to true. This flags the case for manual verification rather than excluding it.

Assign attribution_confidence as follows:

- high: direct evidence of the mechanism in the pharmacist feedback
- moderate: explanation supported but not fully verified
- low: tentative attribution based on limited evidence

If confidence would be low and you cannot identify a clear mechanism, prefer no_clear_reason over forcing a specific category.

For each case, the classifier received the pharmacist’s safety-concern explanation, the system’s recommended dose and interval, the clinician-ordered dose and interval, and the system’s dosing rationale. Autonomy-level labels were excluded from the per-case prompts to avoid anchoring the classifier. The classifier returned a structured response containing: the primary error category (constrained to the seven taxonomy values via JSON schema validation), an attribution confidence level (high, moderate, or low), and a brief evidence statement grounded in the pharmacist feedback and agent rationale.

### Statistical analysis

The dosing event (dosing of an individual patient with an active vancomycin order on a single day of the study) was the unit of analysis. Events were included if all four configurations returned a recommendation and a reference order could be scored (403 of 420 events). All tests were two-sided with *α* = 0.05. No sample size was calculated in advance, and no adjustment was made for repeated events within patients.

#### Dosing agreement (Fig. 2)

Because each dosing event was evaluated by all four system configurations, comparisons between autonomy levels and between guideline conditions used paired tests. Continuous APE was compared using paired *t*-tests (Fig. 2a,b). The proportion of recommendations within the 25% APE tolerance was compared using McNemar’s test on the 2 *×* 2 table of discordant pairs (Fig. 2c,d). Wilson 95% confidence intervals were computed for each condition-specific proportion. Four paired comparisons were conducted in each test family (two autonomy comparisons stratified by guideline condition and two guideline comparisons stratified by autonomy level), and *P* values were adjusted using the Bonferroni method within each family.

#### Safety analysis (Fig. 3)

To test whether completed pharmacist reviews were distributed across the four conditions in proportion to the out-of-tolerance pool, a chi-squared goodness-of-fit test was used, with Monte Carlo simulation of the *P* value when any expected count fell below 5 (Fig. 3a). Safety-concern rates were estimated as the proportion of assessable reviews (Y + N) rated as having a safety concern, with Wilson 95% confidence intervals (Fig. 3b). Because pharmacist reviews were sampled from out-of-tolerance runs and pairing across conditions was incomplete, a logistic regression model was used rather than a paired test: the binary safety-concern rating was regressed on autonomy level and guideline condition, and odds ratios with 95% profile-likelihood confidence intervals were obtained by exponentiating the model coefficients (Fig. 3c). Autonomy-level safety-concern rates pooled across guideline conditions were also compared using Pearson’s chi-squared test (or Fisher’s exact test when any expected cell count fell below 5). To test whether the distribution of LLM-classified error categories differed between autonomy levels, Pearson’s chi-squared test was used on the 2 *×* 6 contingency table of autonomy level by error category, with Monte Carlo simulation of the *P* value (*B* = 10,000) because some expected cell counts were below 5 (Fig. 3d).

Analyses and visualizations were conducted in R (version 4.6.1) using the tidyverse, broom, gt, patchwork and ellmer packages.

## Data Availability

All data produced in the present study are available upon reasonable request to the authors

## Code availability

Code for both configurations will be available at GitHub on publication. The PK engine is a commercial service and is not included.

## Acknowledgements

We thank InsightRx for providing access to its application programming interface for vancomycin dosing. InsightRx had no role in the design of the study, the analysis or interpretation of the data, or the decision to publish. We also thank Stefanie Nguyen, Tran Truong, and Kristine Bielaski, PharmD, for providing independent dosing recommendations and case assessments. Dr. Chen has received research funding support in part by ARPA-H PACT: Physician-AI Collaboration Teaming, NIH/National Institute of Allergy and Infectious Diseases (1R01AI17812101), NIH-NCATS-Clinical & Translational Science Award (UM1TR004921), Stanford Bio-X Interdisciplinary Initiatives Seed Grants Program (IIP) [R12] [JHC], NIH/Center for Undiagnosed Diseases at Stanford (U01 NS134358), Josiah Macy Jr. Foundation (AI in Medical Education), Stanford CARE AI Scholar Fellowship and Stanford Clinical Excellence Research Center.

## Figure legends

**Extended Data Fig. 1.**
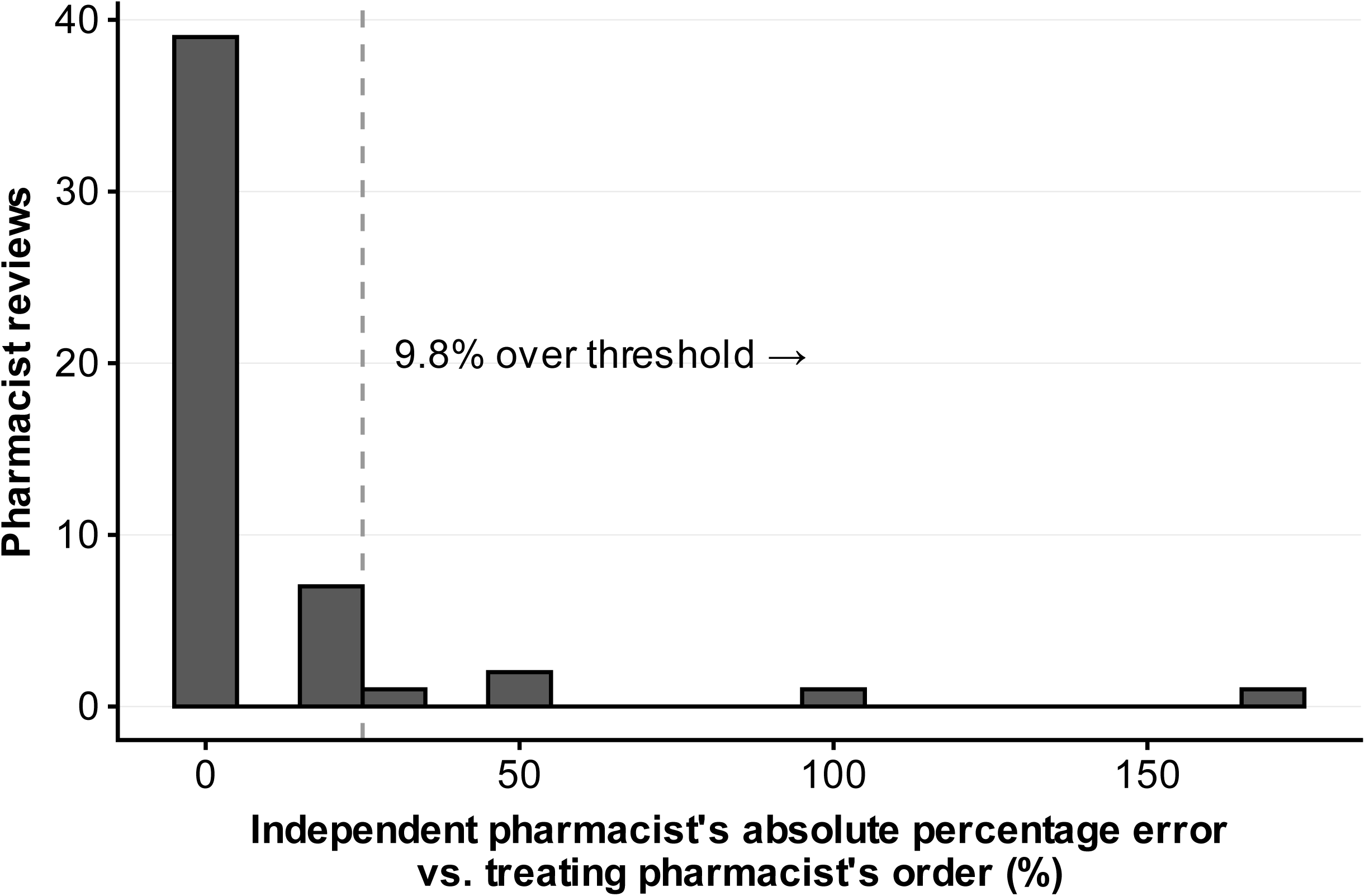
Agreement between independent and treating pharmacists. APE of an independent pharmacist’s recommended total daily dose relative to the treating pharmacist’s order, for 51 blinded reviews of 51 randomly selected dosing events. Bins are 10 percentage points wide; the dashed line marks the 25% tolerance threshold. Forty-six of 51 events (90.2%) were within 25% of the treating pharmacist’s order.

**Extended Data Fig. 2.**
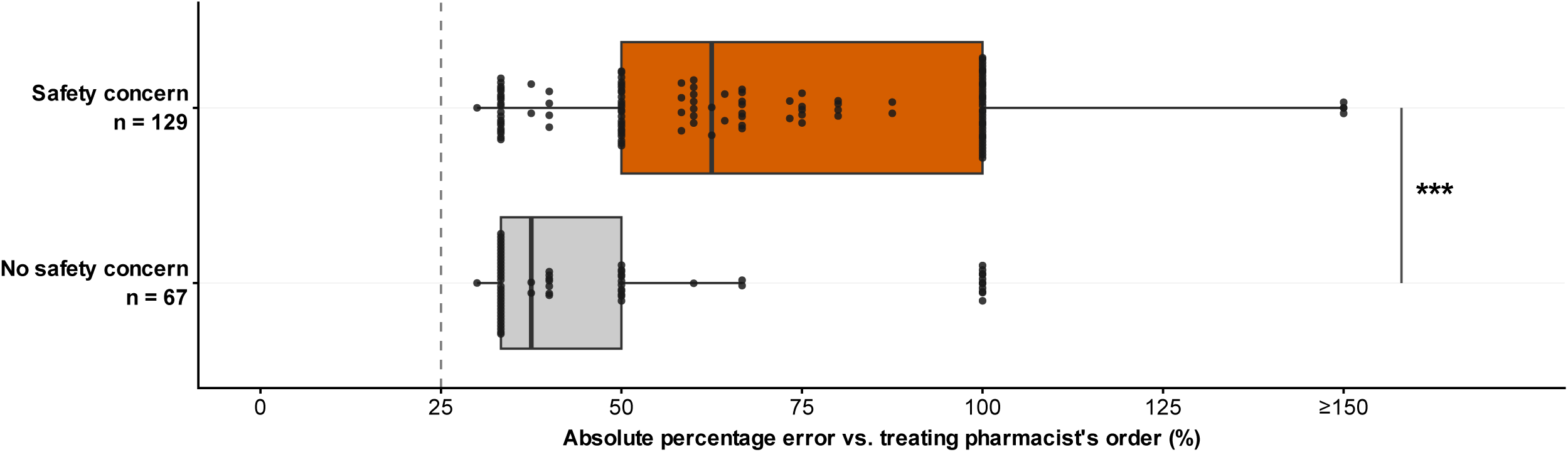
APE by pharmacist safety-concern rating. APE relative to the treating pharmacist’s order for reviewed out-of-tolerance recommendations, by whether the pharmacist identified a safety concern (red-orange; *n* = 129) or not (grey; *n* = 67). Boxes show the median and interquartile range (whiskers, 1.5 *×* interquartile range); points show individual recommendations. Recommendations with a safety concern had higher APE (median, 62.5% (interquartile range, 50.0–100.0%) versus 37.5% (33.3–50.0%); two-sided Wilcoxon rank-sum test, *W* = 2,407.5, *P* = 2.2 *×* 10*^−^*^7^). \*\*\**P <* 0.001. All reviewed recommendations had APE *>* 25% (dashed line); values above 150% are plotted at 150%. Extended Data Fig. 3 breaks the safety-concern group down by error category.

**Extended Data Fig. 3.**
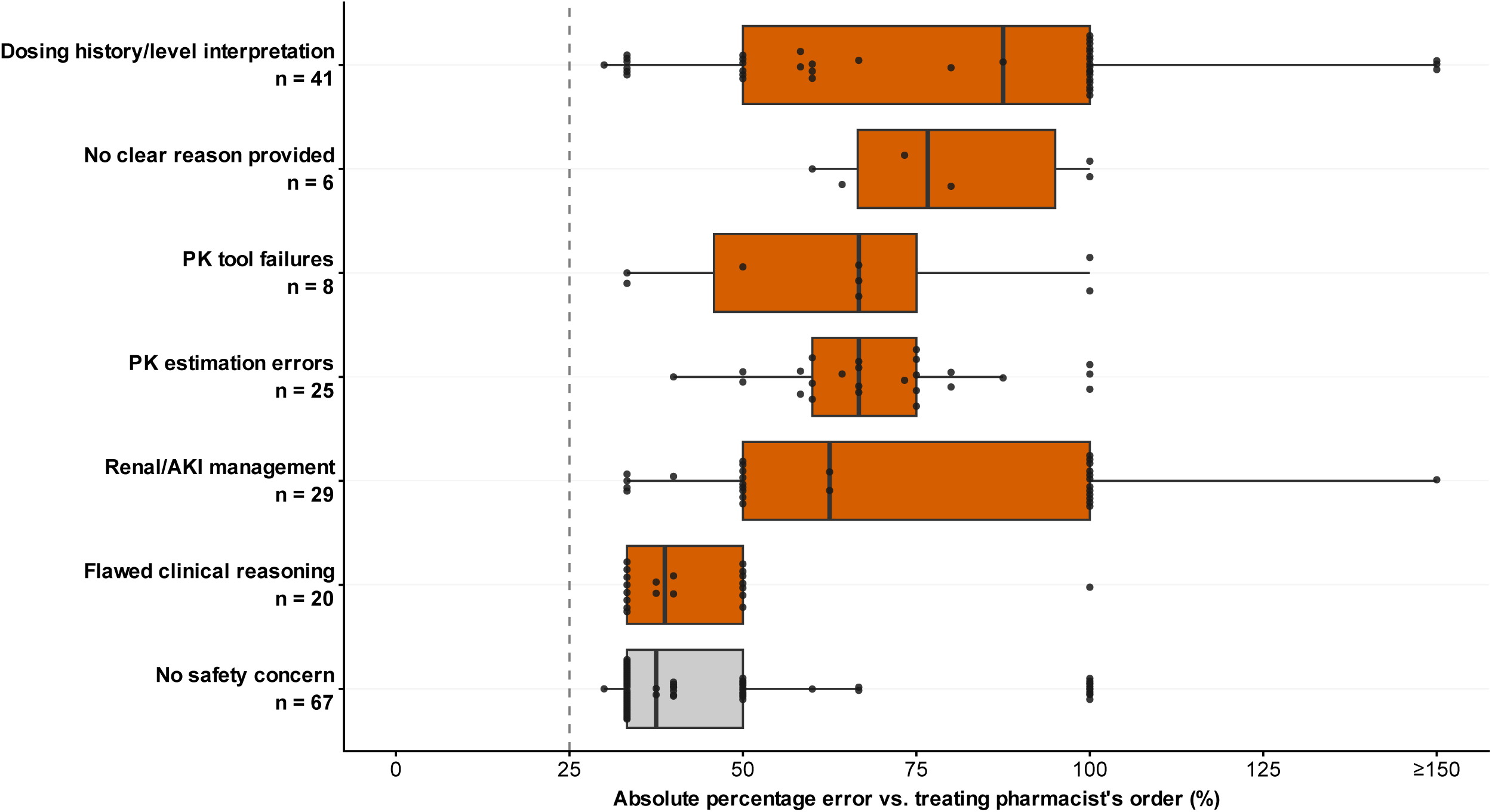
APE by error category. APE relative to the treating pharmacist’s order for the 129 recommendations with a safety concern, by error category (red-orange), with the 67 reviewed recommendations without a safety concern (grey) for reference; group sizes are given on the axis. Categories are ordered by descending median APE. Boxes show the median and interquartile range (whiskers, 1.5 *×* interquartile range); points show individual recommendations. Median APE ranged from 87.5% for dosing history/level interpretation to 38.8% for flawed clinical reasoning, which was similar to recommendations without a safety concern (37.5%). The dashed line marks the 25% review threshold; values above 150% are plotted at 150%.

